# Racism as a Leading Cause of Death: Measuring Excess Deaths in the US

**DOI:** 10.1101/2021.08.30.21262732

**Authors:** Daisy S. Massey, Jeremy S. Faust, Karen B. Dorsey, Yuan Lu, Harlan M. Krumholz

## Abstract

**Background:** Excess death for Black people compared with White people is a measure of health equity. We sought to determine the excess deaths under the age of 65 (<65) for Black people in the United States (US) over the most recent 20-year period. We also compared the excess deaths for Black people with a cause of death that is traditionally reported.

**Methods:** We used the Center of Disease Control (CDC) WONDER’s Multiple Cause of Death 1999-2019 dataset to report age-adjusted mortality rates among non-Hispanic Black (Black) and non-Hispanic White (White) people and to calculate annual age-adjusted <65 excess deaths for Black people from 1999-2019. We measured the difference in mortality rates between Black and White people and the 20-year and 5-year trends using linear regression. We compared age-adjusted <65 excess deaths for Black people to the primary causes of death among <65 Black people in the US.

**Results:** From 1999 to 2019, the age-adjusted mortality rate for Black men was 1,186 per 100,000 and for White men was 921 per 100,000, for a difference of 265 per 100,000. The age-adjusted mortality rate for Black women was 802 per 100,000 and for White women was 664 per 100,000, for a difference of 138 per 100,000. While the gap for men and women is less than it was in 1999, it has been increasing among men since 2014. These differences have led to many Black people dying before age 65. In 1999, there were 22,945 age-adjusted excess deaths among Black women <65 and in 2019 there were 14,444—deaths that would not have occurred had their risks been the same as those of White women. Among Black men, 38,882 age-adjusted excess <65 deaths occurred in 1999 and 25,850 in 2019. When compared to the top 5 causes of deaths among <65 Black people, death related to disparities would be the highest mortality rate among both <65 Black men and women.

**Comment:** In the US, over the recent 20-year period, disparities in mortality rates resulted in between 61,827 excess deaths in 1999 and 40,294 excess deaths in 2019 among <65 Black people. The race-based disparity in the US was the leading cause of death among <65 Black people. Societal commitment and investment in eliminating disparities should be on par with those focused on other leading causes of death such as heart disease and cancer.

## Introduction

In the United States (US), mortality disparities between non-Hispanic Black and non-Hispanic White men and women persist.^1^ One way to understand the impact of this disparity is to determine the number of excess deaths, or deaths that would not have occurred had there been no disparity between Black and White men and women. Age-adjusted differences in deaths, or age-adjusted excess deaths, have not been measured in relation to the racial health disparity between Black and White people in the US in the past 20 years. The most recent estimates are from 2002 and 2004.^2,3^ Additionally, understanding the trend in the mortality disparity is critical to determine whether progress has been made, especially during a period of healthcare expansion.^4^

Using data from CDC WONDER, we measured the age-adjusted excess deaths caused by the health disparity for Black people <65 in the United States over the most recent 20-year and 5-year periods. We compared the number of age-adjusted excess deaths to the top 5 causes of deaths among <65 Black men and women to illuminate the magnitude of age-adjusted excess deaths and to treat systemic racism as its own cause of death, even as it contains parts of all the other causes. In essence, the cause of death among the excess deaths is the social context of being a Black person, since there is no biological reason that Black people should be less likely to live to age 65 than White people.^5-7^ Finally, we show here the age-adjusted mortality rates among Black and White men and women, the differences in rates, and trends in the rates and differences from 1999 to 2019.

## Methods

### Data Source

We used age-adjusted mortality estimates, population estimates, and crude number of deaths from the Center for Disease Control’s WONDER dataset, Multiple Cause of Death, 1999-2019.^8^

### Study Population

Our query parameters included: Not Hispanic or Latino (Hispanic Origin); Black or African American and White (Race); Female and Male (Gender). Race, ethnicity, and gender (henceforth to be referred to as sex) are defined by the person completing the death certificate for everyone represented in the CDC WONDER dataset.^8^

### Statistical Analysis

#### Excess Deaths Due to Mortality Difference

We also calculated age-adjusted annual excess deaths 1999 to 2019 among <65 Black people by sex, defined as deaths that would not have occurred in that year had Black residents experienced the same age-adjusted mortality rates as White residents. We calculated the excess resident deaths by sex and 10-year age subgroup, subtracting from the actual number of deaths reported in the CDC WONDER dataset for each subgroup from those that would have been expected had the rates for Black residents been the same as for White residents. We next found the annual age-adjusted excess deaths and excess death rate per 100,000 people, population estimates from CDC WONDER and weighted averages for age-adjustment from the 2000 United States (US) standard population.^9^ We compared age-adjusted excess death rates to the age-adjusted mortality rates of the top 5 causes of death among <65 Black men and Black women, and using age-adjusted mortality rates by cause of death and groupings from CDC WONDER.

#### Age-Adjusted Mortality Rates

We used and visualized age-adjusted mortality rates per 100,000 people by race (Black or White) and sex (male or female) from CDC WONDER and included the 95% confidence interval (CI) provided. We used rates based on the default intercensal populations for years 2001-2009 (except Infant Age Groups).

#### Mortality Trends

We next estimated the annual trend in each age-adjusted mortality rate and in the difference in mortality rates, by sex, comparing Black and White residents, across the 20-year and 5-year period. First, we fit regression models for each race and sex subgroup with the dependent variable as the age-adjusted mortality rate and the independent variable as time in years. Then, we fit regression models for each sex subgroup with the dependent variable as the difference between age-adjusted mortality rates and the independent variable as time in years. To account for serial correlation of annual outcome rates, we incorporated an autoregressive (AR) error term with a 1-year correlation. We used the coefficient of the time variables to evaluate the slope of each race and sex subgroup’s mortality rate and the mortality difference by sex, from 1999 to 2019 and from 2014 to 2019.

Finally, to determine how the rates and mortality gap changed since 1999 and 2014, we also estimated the total difference in the age-adjusted mortality rate within each sex and race and mortality difference between races within each sex. We measured the total differences between 1999 and 2019 and between 2014 and 2019 by incorporating the standard error of each estimate to construct 95% CIs.

For all analyses, 95% CI excluding 0 was used to determine statistical significance. P value <0.05 was also used to determine statistical significance for annual trends.

All our code is available upon request. All analyses were performed by DM using Stata SE version 16.1 (StataCorp, College Station, TX) and she takes responsibility for the analyses.

## Results

From 1999 to 2019, the number of age-adjusted excess deaths among <65 Black women as compared with White women, deaths that would not have occurred had there been no mortality disparity, declined from 22,945 deaths in 1999 to 14,444 deaths in 2019. Excess deaths among <65 Black men as compared with White men declined from 38,882 deaths in 1999 to 25,851 deaths in 2019.

Compared with the top 5 causes of death among <65 Black women in 1999, age-adjusted excess deaths (139 per 100,000 people) were the highest age-adjusted mortality rate, above malignant neoplasms (83.1 [95% CI: 81.6, 84.7]) and diseases of the heart (66.2 [64.9, 67.6]) (Table 1A, Figure 1). In 2019, age-adjusted excess deaths (74.4) were still the highest age-adjusted mortality rate, above malignant neoplasms (58.7 [57.6, 59.7]) and diseases of the heart (46.1 [45.2, 47.0]). Compared with the top 5 causes of death among <65 Black men, in 1999 age-adjusted excess deaths (252) were the highest age-adjusted mortality rate, above diseases of the heart (128 [126, 130]) and malignant neoplasms (114 [112, 116]) (Table 1B, Figure 1). In 2019, age-adjusted excess deaths (138) were still the highest age-adjusted mortality rate, above diseases of the heart (93.8 [92.4, 95.2]) and malignant neoplasms (63.1 [62.0, 64.2]).

**Table 1A.** Age-Adjusted Mortality Rates (per 100,000) by Top 5 Causes of Death and Excess Death Among <65 Black Women in the US, 1999-2019.

| **Year** | **<65**<br>**Excess**<br>**Deaths** | **Malignant**<br>**neoplasms**<br>**(C00-C97)** | **Diseases of the**<br>**heart (I00-**<br>**I09,I11,I13,I20-**<br>**I51)** | **Accidents**<br>**(unintentional**<br>**injuries) (V01-**<br>**X59,Y85-**<br>**Y86)** | **Cerebrovascular**<br>**diseases (I60-**<br>**I69)** | **Diabetes**<br>**mellitus**<br>**(E10-E14)** |
| --- | --- | --- | --- | --- | --- | --- |
| **1999** | 139 | 83.1 (81.6,<br>84.7) | 66.2 (64.9,<br>67.6) | 17.2 (16.5,<br>17.8) | 16.9 (16.2,<br>17.6) | 15.1 (14.5,<br>15.8) |
| **2000** | 136 | 81.9 (80.4,<br>83.3) | 65.4 (64, 66.7) | 16.7 (16.1,<br>17.4) | 17.4 (16.7,<br>18.1) | 14.3 (13.6,<br>14.9) |
| **2001** | 132 | 80.7 (79.3,<br>82.2) | 64.9 (63.6,<br>66.2) | 17.0 (16.4,<br>17.6) | 16.8 (16.1,<br>17.5) | 14.2 (13.6,<br>14.8) |
| **2002** | 129 | 79.7 (78.3,<br>81.1) | 62.9 (61.7,<br>64.2) | 17.3 (16.6,<br>17.9) | 16.1 (15.5,<br>16.7) | 14.0 (13.4,<br>14.6) |
| **2003** | 127 | 78.6 (77.3,<br>80.0) | 62.3 (61.1,<br>63.5) | 17.2 (16.6,<br>17.9) | 16.2 (15.5,<br>16.8) | 14.2 (13.6,<br>14.8) |
| **2004** | 119 | 76.2 (74.8,<br>77.5) | 58.1 (57.0,<br>59.3) | 17.6 (17.0,<br>18.2) | 15.0 (14.4,<br>15.6) | 13.0 (12.4,<br>13.5) |
| **2005** | 116 | 75.8 (74.5,<br>77.2) | 56.3 (55.2,<br>57.5) | 17.8 (17.1,<br>18.4) | 14.5 (13.9,<br>15.1) | 12.6 (12.1,<br>13.2) |
| **2006** | 110 | 73.3 (72.1,<br>74.6) | 53.4 (52.3,<br>54.5) | 18.1 (17.5,<br>18.7) | 14.0 (13.4,<br>14.5) | 12.4 (11.9,<br>12.9) |
| **2007** | 102 | 71.8 (70.6,<br>73.1) | 50.7 (49.7,<br>51.7) | 17.8 (17.2,<br>18.4) | 13.7 (13.2,<br>14.3) | 12.2 (11.7,<br>12.7) |
| **2008** | 95.8 | 69.7 (68.5,<br>70.9) | 52.0 (50.9,<br>53.0) | 16.1 (15.5,<br>16.7) | 13.4 (12.8,<br>13.9) | 10.9 (10.4,<br>11.4) |
| **2009** | 90.0 | 68.6 (67.4,<br>69.8) | 49.2 (48.2,<br>50.2) | 15.9 (15.4,<br>16.5) | 12.3 (11.8,<br>12.8) | 10.4 (10.0,<br>10.9) |
| **2010** | 84.5 | 68.9 (67.7,<br>70.0) | 47.6 (46.6,<br>48.5) | 15.4 (14.8,<br>16.0) | 12.0 (11.5,<br>12.5) | 10.5 (10,<br>11.0) |
| **2011** | 78.9 | 68.9 (67.7,<br>70.1) | 45.8 (44.8,<br>46.7) | 16.0 (15.4,<br>16.6) | 11.1 (10.6,<br>11.5) | 10.9 (10.5,<br>11.4) |
| **2012** | 75.8 | 66.5 (65.4,<br>67.7) | 45.8 (44.8,<br>46.7) | 16.0 (15.4,<br>16.6) | 11.0 (10.5,<br>11.4) | 10.3 (9.80,<br>10.7) |
| **2013** | 74.9 | 65.3 (64.2,<br>66.4) | 46.3 (45.3,<br>47.2) | 16.8 (16.2,<br>17.3) | 10.4 (9.9, 10.8) | 10.7 (10.3,<br>11.2) |
| **2014** | 73.4 | 65.2 (64.1,<br>66.3) | 46.9 (46.0,<br>47.9) | 17.2 (16.6,<br>17.8) | 10.5 (10.0,<br>10.9) | 10.8 (10.3,<br>11.2) |
| **2015** | 70.8 | 63.8 (62.7,<br>64.9) | 46.6 (45.6,<br>47.5) | 18.2 (17.6,<br>18.8) | 10.5 (10.1,<br>10.9) | 10.8 (10.4,<br>11.3) |
| **2016** | 73.8 | 63.6 (62.6,<br>64.7) | 47.0 (46.1,<br>47.9) | 21.8 (21.1,<br>22.5) | 10.5 (10.1,<br>10.9) | 11.1 (10.6,<br>11.5) |
| **2017** | 70.2 | 60.9 (59.9,<br>62.0) | 46.5 (45.6,<br>47.5) | 23.2 (22.5,<br>23.9) | 10.4 (10.0,<br>10.9) | 11.1 (10.6,<br>11.5) |
| **2018** | 72.3 | 59.5 (58.5, 60.5) | 46.8 (45.9, 47.8) | 23.3 (22.6, 24.0) | 10.0 (9.60, 10.5) | 11.3 (10.9, 11.8) |
| **2019** | 74.0 | 58.7 (57.6, 59.7) | 46.1 (45.2, 47) | 24.6 (23.9, 25.3) | 10.3 (9.90, 10.8) | 11.1 (10.7, 11.6) |

**Data Source:** Age-adjusted mortality rate of top 5 causes of death among <65 Black people, by sex, provided by CDC WONDER Multiple Cause of Death file.

**Table 1B.** Age-Adjusted Mortality Rates (per 100,000) by Top 5 Causes of Death and Excess Death Among <65 Black Men in the US, 1999-2019.

| **Year** | **Excess Deaths** | **Diseases of heart (I00-I09,I11,I13,I20-I51)** | **Malignant neoplasms (C00-C97)** | **Cerebrovascular diseases (I60-I69)** | **Diabetes mellitus (E10-E14)** | **Accidents (unintentional injuries) (V01-X59,Y85-Y86)** |
| --- | --- | --- | --- | --- | --- | --- |
| **1999** | 252 | 128 (126, 130) | 114 (112, 116) | 51.3 (50.1, 52.5) | 38.5 (37.5, 39.5) | 39.6 (38.5, 40.6) |
| **2000** | 239 | 122 (120, 124) | 110 (108, 112) | 49.9 (48.8, 51.1) | 39.9 (38.9, 40.9) | 38.4 (37.3, 39.4) |
| **2001** | 227 | 119 (117, 121) | 105 (103, 107) | 49.0 (47.8, 50.1) | 41.0 (40.0, 42.1) | 37.0 (36.0, 38.1) |
| **2002** | 223 | 119 (117, 121) | 104 (102, 105) | 48.2 (47.1, 49.4) | 41.5 (40.5, 42.5) | 36.1 (35.1, 37.1) |
| **2003** | 219 | 116 (115, 118) | 101 (99.7, 103) | 47.0 (45.9, 48.1) | 42.3 (41.3, 43.3) | 33.9 (32.9, 34.9) |
| **2004** | 206 | 111 (110, 113) | 97.0 (95.4, 98.7) | 47.1 (46.0, 48.2) | 40.3 (39.3, 41.3) | 31.6 (30.7, 32.5) |
| **2005** | 203 | 110 (108, 112) | 94.9 (93.3, 96.5) | 50.0 (48.8, 51.1) | 43.0 (42.0, 44.0) | 30.2 (29.4, 31.1) |
| **2006** | 193 | 106 (104, 107) | 91.2 (89.7, 92.7) | 52.1 (51.0, 53.3) | 44.0 (43.0, 45.0) | 28.1 (27.2, 28.9) |
| **2007** | 181 | 103 (101, 104) | 89.6 (88.1, 91.1) | 48.8 (47.7, 49.8) | 43.0 (42.0, 44.0) | 25.9 (25.1, 26.7) |
| **2008** | 159 | 97.0 (95.4, 98.5) | 87.2 (85.7, 88.6) | 43.2 (42.2, 44.2) | 40.1 (39.2, 41.1) | 22.6 (21.8, 23.3) |
| **2009** | 152 | 96.9 (95.4, 98.4) | 85.4 (84.0, 86.8) | 40.5 (39.6, 41.5) | 37.3 (36.4, 38.2) | 20.5 (19.8, 21.2) |
| **2010** | 137 | 92.6 (91.1, 94.1) | 83.6 (82.2, 84.9) | 39.2 (38.2, 40.1) | 36.7 (35.8, 37.6) | 17.2 (16.5, 17.8) |
| **2011** | 128 | 90.8 (89.4, 92.3) | 80.9 (79.6, 82.2) | 40.6 (39.6, 41.5) | 36.9 (35.9, 37.8) | 14.9 (14.3, 15.5) |
| **2012** | 127 | 91.9 (90.5, 93.3) | 79.2 (77.9, 80.5) | 40.7 (39.8, 41.7) | 38.7 (37.8, 39.7) | 13.1 (12.5, 13.6) |
| **2013** | 125 | 91.2 (89.8, 92.6) | 77.1 (75.9, 78.4) | 42.3 (41.3, 43.2) | 37.3 (36.4, 38.2) | 12.7 (12.1, 13.2) |
| **2014** | 118 | 89.6 (88.2, 91.0) | 75.1 (73.9, 76.4) | 43.7 (42.7, 44.7) | 35.9 (35.0, 36.8) | 11.9 (11.3, 12.4) |
| **2015** | 124 | 90.5 (89.1, 91.9) | 72.7 (71.5, 73.9) | 49.1 (48.0, 50.1) | 42.1 (41.1, 43.0) | 10.8 (10.3, 11.3) |
| **2016** | 127 | 92.6 (91.2, 94.0) | 70.8 (69.6, 72.0) | 57.7 (56.6, 58.8) | 45.2 (44.2, 46.1) | 9.90 (9.40, 10.4) |
| **2017** | 130 | 92.5 (91.1, 93.9) | 66.7 (65.5, 67.8) | 64.4 (63.2, 65.5) | 45.3 (44.3, 46.3) | 8.90 (8.50, 9.40) |
| **2018** | 133 | 94.6 (93.2, 96) | 64.0 (62.9, 65.1) | 64.8 (63.6, 65.9) | 42.3 (41.3, 43.2) | 8.20 (7.80, 8.60) |
| **2019** | 138 | 93.8 (92.4, 95.2) | 63.1 (62.0, 64.2) | 69.4 (68.1, 70.6) | 44.9 (43.9, 45.9) | 7.50 (7.10, 7.90) |

**Data Source:** Age-adjusted mortality rate of top 5 causes of death among <65 Black people, by sex, provided by CDC WONDER Multiple Cause of Death file.

**Table 2.** Trends in US Age-Adjusted Mortality Rates and Mortality Disparity by Race and Sex, 1999-2019 and 2014-2019.

| **Race/ Ethnicity**<br>**by Sex**<br>**Subgroups** | **Annual Rate of**<br>**Change in**<br>**Deaths per**<br>**100,000 (95%**<br>**CI)** | **P Value** | **Total Change**<br>**Since 1999 in**<br>**Deaths per**<br>**100,000 (95%**<br>**CI)** | **Total Change**<br>**Since 2014 in**<br>**Deaths per**<br>**100,000 (95%**<br>**CI)** |
| --- | --- | --- | --- | --- |
| **White Male** | -9.06 (-13.2, -<br>4.93) | 0.00 | -181 (-183, -178) | -7.40 (-9.77, -<br>5.03) |
| **Black Male** | -19.0 (-26.9, -<br>11.1) | 0.00 | -375 (-384, -365) | 14.4 (6.68, 22.1) |
| Gap w/ White<br>Male | -10.1 (-15.7, -<br>4.57) | 0.00 | -194 (-204, -184) | 21.8 (13.7, 29.8) |
| **White Female** | -5.00 (-7.72, -<br>2.28) | 0.00 | -97.3 (-99.1, -<br>95.4) | -8.80 (-10.6, -<br>7.03) |
| **Black Female** | -11.9 (-17.3, -<br>6.42) | 0.00 | -231 (-237, -224) | -15.8 (-20.9, -<br>10.7) |
| Gap w/ White<br>Female | -7.08 (-11.3, -<br>2.86) | 0.00 | -133 (-140, -127) | -7.00 (-12.4, -<br>1.57) |

**Figure 1.**
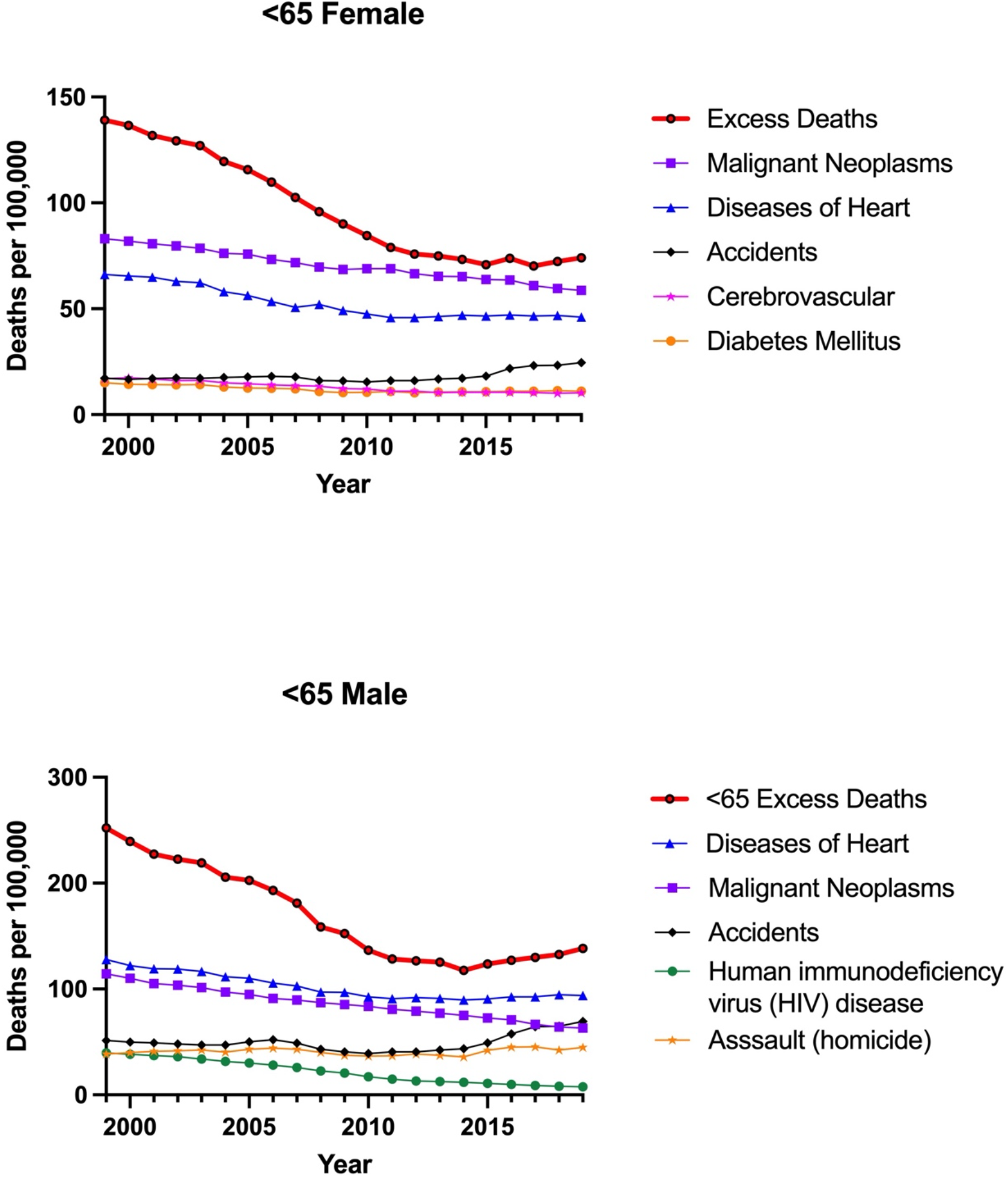
Top 5 Causes of Death and Excess Death Among <65 Black People in the US, By Sex, 1999-2019. **Data Source:** Age-adjusted mortality rates of top 5 causes of death among <65 Black people by sex provided by CDC WONDER Multiple Cause of Death file.

**Figure 2.**
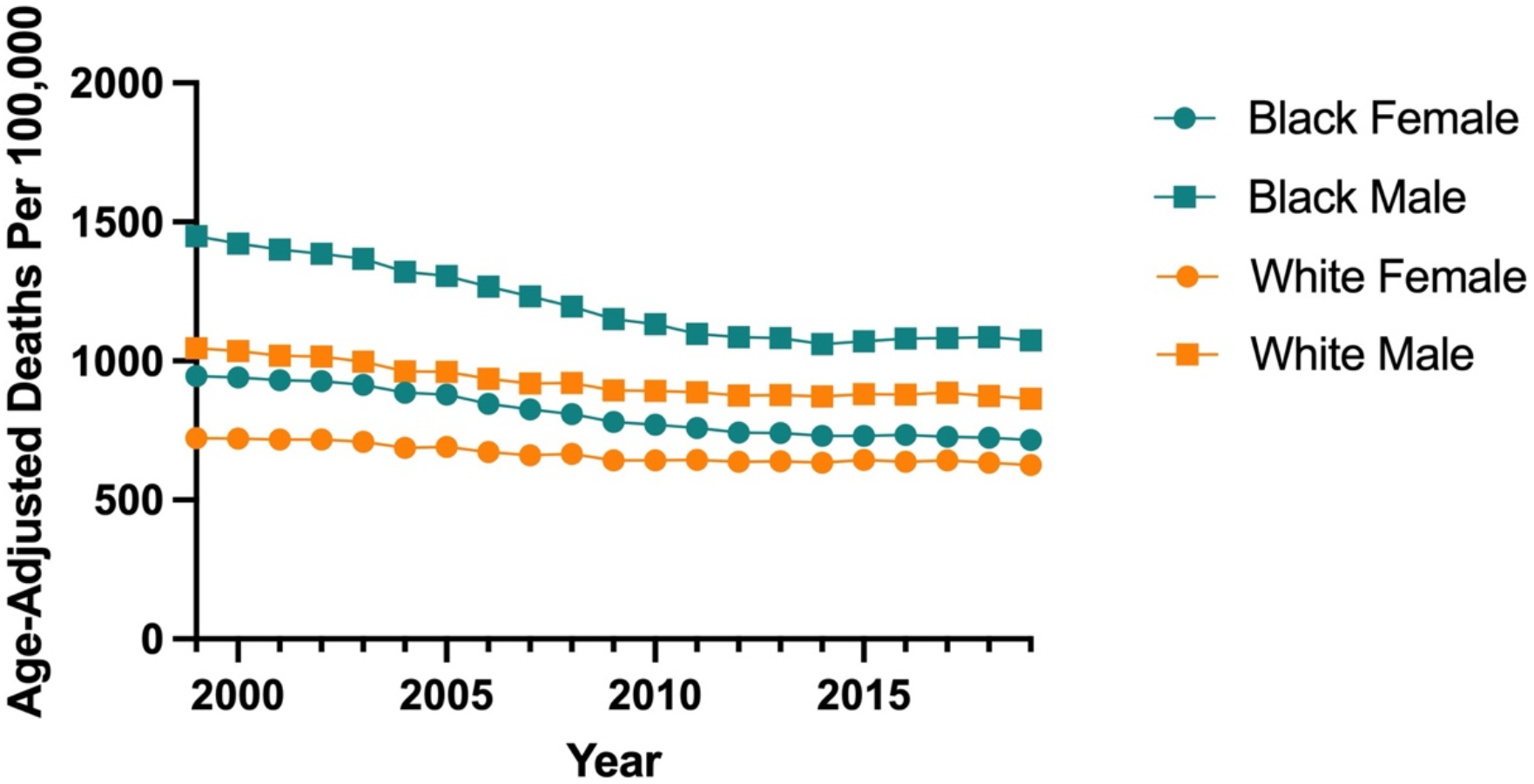
Age-Adjusted Mortality Rates by Sex Among Black and White People in the US, 1999-2019. **Data Source:** Age-adjusted mortality rates by race and sex provided by CDC WONDER Multiple Cause of Death file.

In 1999, the age-adjusted mortality rate per 100,000 people was 946 (941, 951) among Black women, and 722 (721, 724) among White women. By 2019, mortality rates decreased to 715 (712, 719) among Black women and 625 (624, 626) among White women. In 1999, the age-adjusted mortality rate was 1,450 (1,440, 1,460) among Black men and 1,050 (1,040, 1,050) among White men. In 2019, the age-adjusted mortality rate was 1,070 (1,070, 1,080) among Black men and 865 (863, 867) among White men. From 1999 to 2019, the average mortality rate per 100,000 was 802 (801, 803) among Black women, 664 (663, 664) among White women, 1,186 (1,184; 1,187) among Black men, and 921 (921, 922) among White men.

From 1999 to 2019, age-adjusted mortality rates per 100,000 people decreased among Black women (−231 [-237, -224]) and White women (−97.3 [-99.1, -95.4]), and the gap between Black and White female mortality rates decreased (−133 [-140, -127]). Age-adjusted mortality rates per 100,000 decreased among Black men (−375 [-384, -365]) and White men (−181 [-183, - 178]), and the gap between Black and White male mortality rates also decreased (−194 [-204, - 184]).

From 2014 to 2019, the age-adjusted mortality rates decreased among Black women (−15.8 [-20.9, -10.7]), White women (−8.80 [-10.6, -7.03]), and White men (−7.40 [-9.77, -5.03]), but increased among Black men (14.4 [6.68, 22.1]). The gap between Black and White female mortality rates decreased (−7.00 [-12.4, -1.57]), but the gap between Black and White male mortality rates increased (21.8 [13.7, 29.8]).

## Discussion

In sheer numbers, the age-adjusted excess deaths associated with being identified as a Black person are the leading cause of death among <65 Black men and women.^10^ The age-adjusted excess death rate would be the highest mortality rate among Black women and Black men. These differences led to 61,826 Black men and women dying in 1999 before reaching age 65, and to 40,295 dying in 2019.

Excess deaths represent the difference between the number of deaths among Black residents that occurred and the number of deaths that would have occurred had the mortality rate been the same as that among White people. Excess deaths represent the timing of deaths—each person will die once in a lifetime, but excess deaths capture the disparity in early deaths. The excess deaths associated with race can be understood as a toll that results from racism in the United States.^5-7^ There is no biological reason, independent of social context, that Black people should have a higher mortality than White people. Our data support this. First, almost one third of the disparity between Black and White women decreased over the past 20 years, indicating that, while medicine and public health improved among all people, it improved (relatively) more among Black women based on 1999 rates. Second, while the disparity between Black and White people has decreased among women since 2014, it has increased among men over the same time period, indicating further that biology is not the cause. Thus, we have put excess deaths by race in context by comparing the toll with causes of death by disease category.

Our study adds importantly to the prior literature. Our study adds importantly to the prior literature. It has been previously estimated that 2,687,051 excess deaths occurred from 1970 to 2004, a 35-year period.^3^ It has also been estimated based on data from 2002, that closing the mortality gap between Black and White people in the US would eliminate 83,570 excess deaths.^2^ Our paper adds a more recent comprehensive update and a focus on early (<65) excess death, finding that over the past 20 years, between about 40,000 and 60,000 Black men and women have died each year due to the mortality disparity. In addition, we add a comparison between the number of excess deaths and the number of Black people who die from the top 5 causes of death to show the full impact of the mortality disparity.

Prior to 2014, it has been shown that the mortality disparity narrowed. From 2000 to 2014, mortality among Black men decreased and the overall disparity between Black and White men narrowed, although not within all causes.^11^ In both sexes, the disparity in overall mortality between Black and White people decreased from 33% in 1999 to 16% in 2015.^12^ However, a more recent report warned that the health disparity in life expectancy, which had been narrowing since 2000, may have begun to widen in 2015 and 2016, but it was too early to identify a certain trend at that time.^13^ Our finding shows that the disparity in mortality rates between Black and White men has widened from 2014 to 2019, but has continued to decrease between Black and White women.

Our finding that excess deaths among <65 Black people are the leading cause of death for Black men and women is a sobering reason to report excess deaths and to treat the mortality disparity conceptually as a cause of death. Although excess deaths are often determined *by* causes of death as opposed to *compared to* causes of death, showing the measure alongside cause of death illuminates the full cost to Black lives. Further, this approach reflects the fact that systemic racism causes health disparities and, consequently, excess deaths.^6^ The deaths captured by the rate of excess deaths and the mortality rate by cause of death overlap, but when excess deaths are isolated, it emphasizes the pervasiveness of the mortality gap and health disparities, rather than treating both as merely clinical or disease-specific issues.^14^

Our study was limited in several ways. First, death certificates, from which CDC WONDER estimates are derived, are known to include inaccurcies in race, ethnicity, and cause of death.^15,16^ However, race and Hispanic origin has been found to be most accurate for non-Hispanic Black and White decedent’s death certificates, in that it more often matched self-reporting in US Census data.^17^ Further, cause of death inaccuracies are most often within the ICD-10 code level, rather than in the ICD-10 grouping that we used.^18,19^

## Conclusion

Excess deaths among <65 Black people in the US led to over 40,000 early deaths each year that would not have occurred otherwise. As compared to the top 5 causes of death, the age-adjusted excess death rate would be as large as the leading cause of death among <65 Black men and women. With the mortality disparity between Black and White men increasing since 2014, excess deaths due to the mortality disparity should be treated as urgently as other leading causes of death such as heart disease and cancer.

## Data Availability

All data are available upon request. All data used in analyses were downloaded from CDC Wonder.

## References

1. National Center for Health S. Health, United States. In: Health, United States, 2015: With Special Feature on Racial and Ethnic Health Disparities. Hyattsville (MD): National Center for Health Statistics (US); 2016.

2. Satcher D, Fryer GE, McCann J, Troutman A, Woolf SH, Rust G. What If We Were Equal? A Comparison Of The Black-White Mortality Gap In 1960 And 2000. Health Affair. 2005;24(2):459–464.

3. Rodriguez JM, Geronimus AT, Bound J, Dorling D. Black lives matter: Differential mortality and the racial composition of the U.S. electorate, 1970–2004. Soc Sci Med. 2015;136-137:193–199.

4. Mazurenko O, Balio CP, Agarwal R, Carroll AE, Menachemi N. The Effects Of Medicaid Expansion Under The ACA: A Systematic Review. Health Aff (Millwood). 2018;37(6):944–950.

5. Churchwell K, Elkind Mitchell SV, Benjamin Regina M, et al. Call to Action: Structural Racism as a Fundamental Driver of Health Disparities: A Presidential Advisory From the American Heart Association. Circulation. 2020;142(24):e454–e468.

6. Bailey ZD, Krieger N, Agénor M, Graves J, Linos N, Bassett MT. Structural racism and health inequities in the USA: evidence and interventions. Lancet. 2017;389(10077):1453–1463.

7. Cogburn CD. Culture, Race, and Health: Implications for Racial Inequities and Population Health. The Milbank Quarterly. 2019;97(3):736–761.

8. Multiple Cause of Death 1999-2019. In: Centers for Disease Control and Prevention NCfHS, ed. CDC WONDER Online Database 2020.

9. Klein RS, C. Age Adjustment Using the 2000 Projected U.S. Population. Healthy People 2010. 2001.

10. Centers for Disease Control and Prevention NCfHSMCoD-oCWOD, released in 2020. Data are from the Multiple Cause of Death Files, 1999-2019, as compiled from data provided by the 57 vital statistics jurisdictions through the Vital Statistics Cooperative Program. In.

11. Pathak EB. Mortality Among Black Men in the USA. J Racial Ethn Health Disparities. 2018;5(1):50–61.

12. Cunningham TJ, Croft JB, Liu Y, Lu H, Eke PI, Giles WH. Vital Signs: Racial Disparities in Age-Specific Mortality Among Blacks or African Americans - United States, 1999-2015. MMWR Morb Mortal Wkly Rep. 2017;66(17):444–456.

13. Bilal U, Diez-Roux AV. Troubling Trends in Health Disparities. N Engl J Med. 2018;378(16):1557–1558.

14. Airhihenbuwa CO, Liburd L. Eliminating Health Disparities in the African American Population: The Interface of Culture, Gender, and Power. Health Education & Behavior. 2006;33(4):488–501.

15. Arias EH, M; Hakes, JK. The validity of race and Hispanic-origin reporting on death certificates in the United States: An update. Hyattsville, Maryland: National Center for Health Statistics; 2016.

16. Hoffman RA, Venugopalan J, Qu L, Wu H, Wang MD. Improving Validity of Cause of Death on Death Certificates. Proceedings of the 2018 ACM International Conference on Bioinformatics, Computational Biology, and Health Informatics; 2018; Washington, DC, USA.

17. Arias ES, WS; Eschbach, K; et al. The validity of race and Hispanic origin reporting on death certificates in the United States. Hyattsville, MD: National Center for Health Statistics; 2008.

18. Falci L, Lee Argov EJ, Van Wye G, Plitt M, Soto A, Huynh M. Examination of Cause-of-Death Data Quality Among New York City Deaths Due to Cancer, Pneumonia, or Diabetes From 2010 to 2014. Am J Epidemiol. 2018;187(1):144–152.

19. Moghanaki D, Howard LE, De Hoedt A, et al. Validity of the National Death Index to ascertain the date and cause of death in men having undergone prostatectomy for prostate cancer. Prostate Cancer and Prostatic Diseases. 2019;22(4):633–635.

